# External validation and head-to-head comparison of Eclipse-PRISM and Johns Hopkins ACG risk scores for predicting emergency admissions in an English older population

**DOI:** 10.64898/2026.06.23.26356325

**Authors:** Dahai Yu, Tim Winters, Anastasija Pinchuk

**Author notes:** **Correspondence:** Dr Dahai Yu, School of Medicine, Keele University, Keele, Staffordshire, the United Kingdom, ST5 5BG.

## Abstract

**Background:** Predictive risk stratification tools are widely used to support proactive care for older adults, yet head-to-head external validation within local English health systems remains limited. Eclipse-PRISM implemented in UK primary care settings, while the Johns Hopkins Adjusted Clinical Groups (ACG) system provides risk scores derived from diagnosis groupings and healthcare utilisation data.

**Aim:** To externally validate and compare PRISM and Johns Hopkins ACG scores for predicting emergency hospital admission among older adults in an English integrated care system.

**Methods:** We conducted a retrospective cohort study in the Norfolk and Waveney Integrated Care System. Individuals aged ≥75 years at the index date with valid PRISM and ACG emergency admission risk scores and linkage to hospital activity data were included. The primary outcome was ≥1 emergency hospital admission within 1 month. Discrimination was assessed using the area under the receiver operating characteristic curve (AUC), with paired AUCs compared using DeLong’s test. Calibration was evaluated using calibration plots and quantified using calibration intercept and slope from logistic recalibration models. Overall accuracy was summarised using the Brier score. Clinical utility was assessed using decision curve analysis (DCA).

**Results:** The cohort included 114,407 patients aged ≥75 years; 2,136 (1.87%) had ≥1 emergency admission within 1 month. ePRISM showed higher discrimination than Johns Hopkins ACG (AUC 0.860 [95% CI 0.852 to 0.867] vs 0.739 [95% CI 0.728 to 0.749]; ΔAUC 0.121 [95% CI 0.111 to 0.130]; DeLong p<0.0001), with consistent differences across age and sex subgroups. Calibration differed materially: ePRISM showed closer agreement between predicted and observed risks, whereas Johns Hopkins ACG systematically overpredicted risk across much of the range. Brier scores favoured ePRISM (0.017 [95% CI 0.017 to 0.018] vs 0.051 [95% CI 0.050 to 0.051]). In DCA, ePRISM provided higher net benefit across clinically plausible thresholds, while Johns Hopkins ACG showed lower or negative net benefit across much of the threshold range.

**Conclusions:** In this English older population, ePRISM demonstrated higher discrimination and more favourable apparent calibration, overall accuracy and decision-analytic performance for predicting 1-month emergency admission than Johns Hopkins ACG. Model selection for short-term risk stratification should therefore consider calibration and clinical utility alongside discrimination, with local validation and recalibration where appropriate before implementation.

## INTRODUCTION

Emergency hospital admissions among older adults are common, costly and frequently associated with functional decline, institutionalisation and mortality [1]. In England, demographic ageing and increasing multimorbidity have contributed to sustained pressure on unplanned care, prompting policy initiatives that encourage Integrated Care Systems (ICSs) to use risk stratification tools to identify individuals at high risk of emergency admission and to target proactive interventions [2].

A range of emergency admission risk stratification (EARS) tools are now deployed across UK primary care. These include proprietary models such as the Johns Hopkins Adjusted Clinical Groups (ACG) system, and locally implemented tools such as PRISM (Predictive Risk Stratification Model) [3]. The Johns Hopkins ACG system uses diagnostic codes, prescriptions and patterns of health-care use to generate morbidity groupings and risk scores that can predict outcomes such as emergency admission and high cost [4]. ePRISM, embedded in the Eclipse prescribing and risk platform, uses large numbers of primary-care and prescribing variables, fitted using machine-learning methods, to produce patient-level emergency admission risk scores [5, 6].

Although these tools are widely promoted to support “case finding” and personalised care, the evidence that their introduction reduces emergency admissions or improves outcomes is mixed. Observational work in English primary care has shown that EARS tools are reasonably accurate in ranking risk, but their clinical usefulness is uncertain and often limited by poor implementation and unclear intervention pathways [3, 7].

Historically, evaluations have emphasised discrimination (e.g. the C-statistic), with less attention to calibration or decision-analytic measures. Recent extensions to the TRIPOD statement, including TRIPOD+AI, stress that calibration and clinical utility are essential components of model evaluation, particularly for machine-learning-based tools [8]. A model with excellent discrimination but poor calibration may produce systematically biased risk estimates, leading to inappropriate thresholds and mis-targeted interventions [9]. Decision curve analysis (DCA) provides a framework to quantify net benefit across risk thresholds and is increasingly recommended alongside discrimination and calibration [10].

In the current study, we report an external validation and head-to-head comparison of the ePRISM score and the Johns Hopkins ACG emergency admission risk score for older adults (≥75 years) in the Norfolk and Waveney Integrated Care System in England. We compare discrimination, calibration, overall accuracy and decision-analytic net benefit to inform local commissioning and highlight broader lessons for selecting risk models in older populations.

## METHODS

### Study design and data setting

We undertook a retrospective cohort study using routinely collected data from general practices within the Norfolk and Waveney Integrated Care System (ICS). In April 2025 Norfolk and Waveney had 105 GP practices and served a registered population of about 1,100,000 [11]. The ICB is the third oldest in England with about 142,500 people aged 75 and over [12] and has a higher-than-average proportion of the total population with long term conditions [13]. Participating practices used the Eclipse platform, which includes pre-computed ePRISM scores, risk dashboards and prescribing data. Primary-care data were linked, via standard NHS processes in Data Hub to hospital activity to obtain information on emergency admissions.

### Study population

Eligible participants were those who (1) were registered with a participating general practice in Norfolk and Waveney ICS; (2) were aged ≥75 years at the index date; (3) had a valid ePRISM risk score recorded; (4) had a valid Johns Hopkins ACG emergency admission risk score recorded; and (5) were successfully linked to HES. After applying these criteria, 114,407 patients formed the analysis cohort (**Figure 1**). The use of this annoymised database in this manner was approved by the South West - Exeter Research Ethics Committee (REC reference: 17/SW/0001).

**Figure 1.**
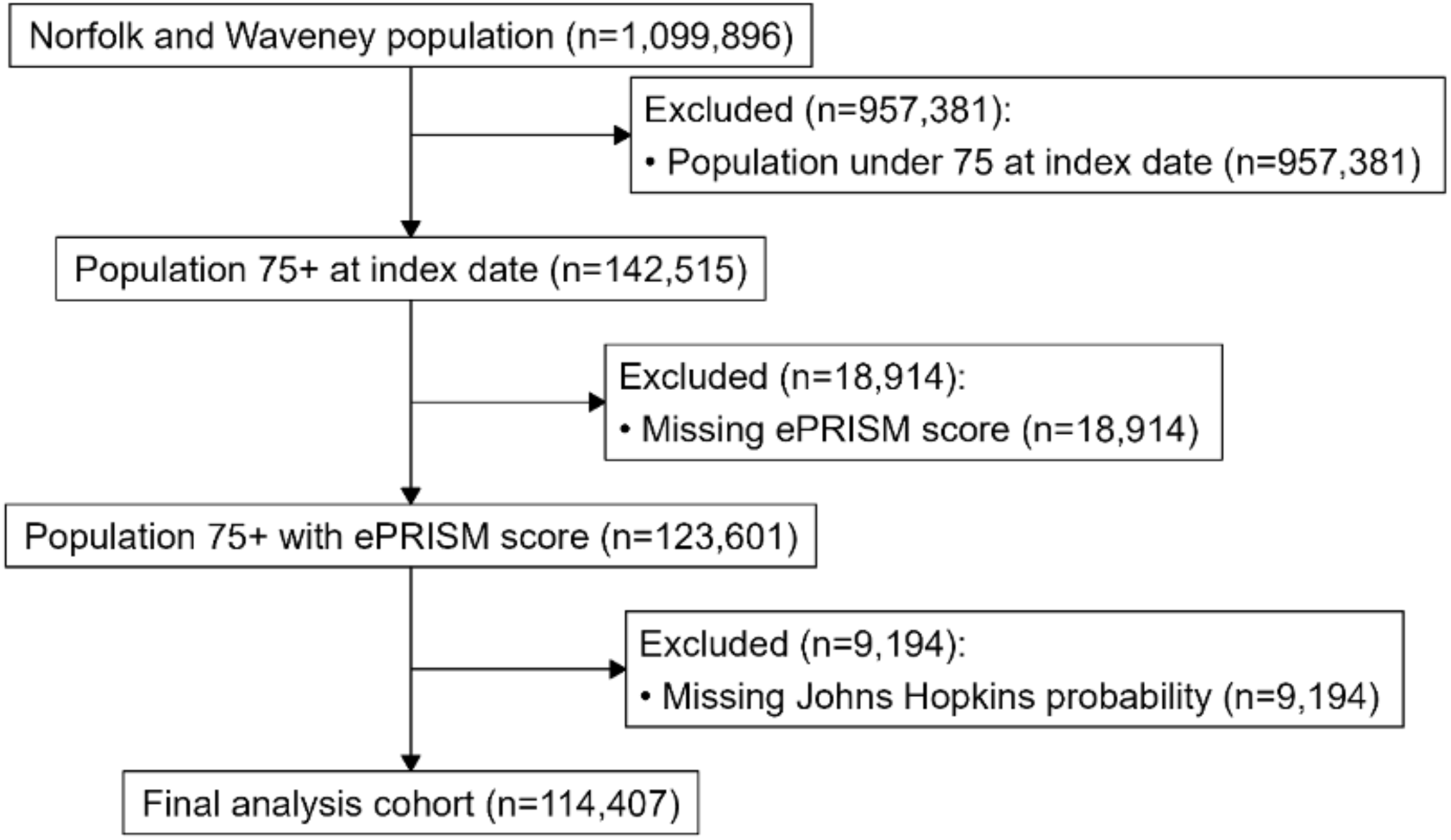
Study cohort selection flow diagram.

### Risk scores

#### ePRISM risk score

ePRISM is a proprietary risk prediction model implemented within Eclipse. It incorporates age, sex, long-term conditions, medication history, diagnostic codes and previous health-care utilisation, with a large number of candidate predictors (exact specification and implementation details to be confirmed locally). For this study, we used the existing ePRISM score as recorded in Eclipse at the index date. The ePRISM score was generated 06/03/2025.

#### Johns Hopkins ACG emergency admission risk score

The Johns Hopkins ACG system is a patient-level case-mix and risk stratification system that categorises diagnoses and other data into morbidity groups and produces risk estimates for outcomes such as emergency department (ED) visits and admissions [14]. In this study, the Johns Hopkins ACG emergency admission risk probability was provided to the ICB as a monthly snap shot as part of the ICB data sharing contract with Arden GEM CSU. The Johns Hopkins emergency admission risk probability is derived using age, gender, disease burden and disease markers, medication, previous utilization and other applicable population markers [15]. The 2020 UK NHS recalibration exercise indicated that the emergency admission AUC statistic was 0.768. The monthly snap shot used in this analysis was generated 31/03/2025.

#### Outcome

The primary outcome was emergency hospital admission over 1 month, defined as ≥1 unplanned (emergency) admission identified in Data Hub between the index date and 1 month thereafter (follow-up window: 01/04/2025 to 30/04/2025). Patients with at least one such admission were coded as cases; all others were non-cases. Emergency admissions were ascertained from the Data Hub linked dataset using the admission method field to identify unplanned/emergency admissions (Admission Method Code in 21, 22, 23, 24, 25, 2A, 2B, 2D, 28) and only admissions occurring within the follow-up window were counted. Where multiple emergency admissions occurred within follow-up, the primary outcome remained binary (any vs none).

## Statistical analysis

All analyses were conducted on the same cohort so that performance comparisons between PRISM and the Johns Hopkins ACG score were paired within individuals.

### Discrimination

We calculated the **C-statistic** (area under the receiver operating characteristic curve, AUC) with 95% confidence intervals (CIs) for each model. The C-statistic can be interpreted as the probability that, for a randomly selected pair of patients (one with and one without an emergency admission), the patient who was admitted has the higher predicted risk.

To compare AUCs, we used DeLong’s non-parametric test for correlated ROC curves [16]. The test statistic is

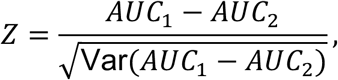

which is approximately standard normal under the null hypothesis that the two C-statistics are equal. We also calculated AUCs stratified by age band (75–79, 80–84, ≥85 years) and sex.

### Calibration

Following TRIPOD and TRIPOD+AI guidance [17, 8], we assessed calibration using multiple complementary approaches. Calibration was examined graphically using calibration plots by grouping predicted risks into deciles and, within each decile, estimating the observed event rate and plotting observed versus predicted risks against a 45° reference line. We also quantified calibration using the calibration intercept and slope by fitting logistic regression models with the observed outcome as the dependent variable and the logit of the predicted probability as the sole predictor, where the ideal intercept is 0 and the ideal slope is 1 [18]. We additionally computed the Hosmer–Lemeshow χ^2^ statistic using deciles of predicted risk; given the very large sample size, this was interpreted descriptively as an indicator of the magnitude of miscalibration rather than relying on statistical significance alone [19]. Overall accuracy was summarised using the Brier score, defined as the mean squared error between predicted probabilities and observed binary outcomes (range 0–1, lower is better) [20].

Decision curve analysis: To evaluate clinical utility, we used decision curve analysis (DCA), which quantifies net benefit across a range of risk thresholds compared with “treat all” and “treat none” strategies [10]. Net benefit was calculated as:

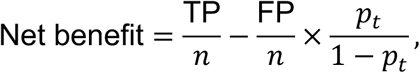

where TP and FP are numbers of true and false positives, *n* is the sample size, and *p_t_* is the chosen threshold probability. We examined thresholds across a plausible range for targeting enhanced case management in older adults (for example 2–20%).

Records were only included if the patient had age, sex, ePRISM score, Johns Hopkins emergency admission risk probability and were linkable to hospital activity in Data Hub. No records were missing for the essential analysis variables, while missing/not-recorded categories were retained for descriptive predictor domains and no imputation was performed..

All analyses were performed using R 4.6.0 (“Because it was There”) statistical software within IDE R Studio 2026.04.0 Build 526.

## RESULTS

### Cohort characteristics

The 114,407 patients aged ≥75 years had a high burden of comorbidity and previous emergency utilisation (**Table 1**). The mean age was 81.5 years (SD 5.3). Women formed a slight majority (55%). Most patients were recorded as White (81%), with 0.3% recorded as non-White and 18.2% not recorded. Ten percent lived in the most deprived communities (top 20% of Index of Multiple Deprivation). Patient need group classifications were most commonly multi-morbidity medium complexity (33%) and multi-morbidity low complexity (21%), with 10% classified as frailty and 10% as multi-morbidity high complexity. Blood pressure categories were most commonly hypertension stage 1 (38%) and prehypertension (37%). Over the 1-month follow-up, 2,136 patients experienced ≥1 emergency admission (1.87%, 95% CI 1.79–1.95%).

**Table 1.** Baseline characteristics of the cohort by emergency admission within 1 month.

|  | Emergency admission - No<br>(N=112271) | Emergency Admission - Yes<br>(N=2136) | Overall<br>(N=114407) |
| --- | --- | --- | --- |
| **Sex** |  |  |  |
| F | 61653 (54.9%) | 1130 (52.9%) | 62783 (54.9%) |
| M | 50618 (45.1%) | 1006 (47.1%) | 51624 (45.1%) |
| **Age** |  |  |  |
| Mean (SD) | 81.5 (5.30) | 83.4 (5.77) | 81.5 (5.31) |
| Median [Min, Max] | 80.0 [75.0, 110] | 83.0 [75.0, 102] | 80.0 [75.0, 110] |
| **Ethnic Group** |  |  |  |
| Any other ethnic group | 106 (0.1%) | 0 (0%) | 106 (0.1%) |
| Asian or Asian British | 134 (0.1%) | 1 (0.0%) | 135 (0.1%) |
| Black or Black British | 35 (0.0%) | 1 (0.0%) | 36 (0.0%) |
| Chinese | 77 (0.1%) | 0 (0%) | 77 (0.1%) |
| Mixed | 48 (0.0%) | 1 (0.0%) | 49 (0.0%) |
| Not recorded | 20343 (18.1%) | 528 (24.7%) | 20871 (18.2%) |
| Whit British | 90955 (81.0%) | 1596 (74.7%) | 92551 (80.9%) |
| White Other | 573 (0.5%) | 9 (0.4%) | 582 (0.5%) |
| **Age Band** |  |  |  |
| 75-79 | 50331 (44.8%) | 648 (30.3%) | 50979 (44.6%) |
| 80-84 | 32496 (28.9%) | 634 (29.7%) | 33130 (29.0%) |
| 85+ | 29444 (26.2%) | 854 (40.0%) | 30298 (26.5%) |
| **Core 20 most deprived communities** |  |  |  |
| No | 101598 (90.5%) | 1863 (87.2%) | 103461 (90.4%) |
| Yes | 10673 (9.5%) | 273 (12.8%) | 10946 (9.6%) |
| **Johns Hopkins Patient Need Group** |  |  |  |
| 01 Non-User | 110 (0.1%) | 0 (0%) | 110 (0.1%) |
| 03 Low Need Adult | 10128 (9.0%) | 41 (1.9%) | 10169 (8.9%) |
| 04 Multi-Morbidity Low Complexity | 23357 (20.8%) | 155 (7.3%) | 23512 (20.6%) |
| 05 Multi-Morbidity Medium Complexity | 36982 (32.9%) | 500 (23.4%) | 37482 (32.8%) |
| 08 Dominant Psychiatric/Behavioural Condition | 2951 (2.6%) | 49 (2.3%) | 3000 (2.6%) |
| 09 Dominant Major Chronic Condition | 16589 (14.8%) | 331 (15.5%) | 16920 (14.8%) |
| 10 Multi-Morbidity High Complexity | 11035 (9.8%) | 468 (21.9%) | 11503 (10.1%) |
| 11 Frailty | 11119 (9.9%) | 592 (27.7%) | 11711 (10.2%) |
| **Chronic Condition Count** |  |  |  |
| Mean (SD) | 5.33 (3.70) | 8.37 (4.61) | 5.39 (3.74) |
| Median [Min, Max] | 5.00 [0, 34.0] | 8.00 [0, 27.0] | 5.00 [0, 34.0] |
| **Alcohol Category** |  |  |  |
| Non Drinker | 43372 (38.6%) | 1016 (47.6%) | 44388 (38.8%) |
| Low Risk | 60317 (53.7%) | 968 (45.3%) | 61285 (53.6%) |
| Increasing Risk | 4248 (3.8%) | 64 (3.0%) | 4312 (3.8%) |
| High Risk | 1089 (1.0%) | 17 (0.8%) | 1106 (1.0%) |
| Alcohol Dependency | 378 (0.3%) | 8 (0.4%) | 386 (0.3%) |
| Unknown | 633 (0.6%) | 17 (0.8%) | 650 (0.6%) |
| Missing | 2234 (2.0%) | 46 (2.2%) | 2280 (2.0%) |
| **BMI Category** |  |  |  |
| Underweight | 2682 (2.4%) | 92 (4.3%) | 2774 (2.4%) |
| Healthy weight | 37851 (33.7%) | 760 (35.6%) | 38611 (33.7%) |
| Overweight | 42861 (38.2%) | 718 (33.6%) | 43579 (38.1%) |
| Obesity | 25369 (22.6%) | 493 (23.1%) | 25862 (22.6%) |
| Severe obesity | 2105 (1.9%) | 60 (2.8%) | 2165 (1.9%) |
| Missing | 1403 (1.2%) | 13 (0.6%) | 1416 (1.2%) |
| **Smoking Status** |  |  |  |
| Non-Smoker | 58519 (52.1%) | 989 (46.3%) | 59508 (52.0%) |
| Ex-Smoker | 47720 (42.5%) | 1014 (47.5%) | 48734 (42.6%) |
| Smoker | 5560 (5.0%) | 126 (5.9%) | 5686 (5.0%) |
| Declined | 150 (0.1%) | 5 (0.2%) | 155 (0.1%) |
| Missing | 322 (0.3%) | 2 (0.1%) | 324 (0.3%) |
| **Total Cholesterol Category** |  |  |  |
| Within range | 75134 (66.9%) | 1542 (72.2%) | 76676 (67.0%) |
| Above range | 34273 (30.5%) | 568 (26.6%) | 34841 (30.5%) |
| Missing | 2864 (2.6%) | 26 (1.2%) | 2890 (2.5%) |
| **HbA1c Category** |  |  |  |
| Normal | 57220 (51.0%) | 1018 (47.7%) | 58238 (50.9%) |
| Prediabetes | 31538 (28.1%) | 590 (27.6%) | 32128 (28.1%) |
| Diabetes | 17323 (15.4%) | 470 (22.0%) | 17793 (15.6%) |
| Missing | 6190 (5.5%) | 58 (2.7%) | 6248 (5.5%) |
| **Blood Pressure Category** |  |  |  |
| Hypotension | 1821 (1.6%) | 78 (3.7%) | 1899 (1.7%) |
| Normal | 10790 (9.6%) | 286 (13.4%) | 11076 (9.7%) |
| Pre-hypertensive | 41397 (36.9%) | 744 (34.8%) | 42141 (36.8%) |
| Hypertension Stage 1 | 43280 (38.5%) | 689 (32.3%) | 43969 (38.4%) |
| Hypertension Stage 2 | 13552 (12.1%) | 306 (14.3%) | 13858 (12.1%) |
| Hypertension Stage 3 (Crisis) | 971 (0.9%) | 32 (1.5%) | 1003 (0.9%) |
| Missing | 460 (0.4%) | 1 (0.0%) | 461 (0.4%) |
| **Em Adm April 2024 To March 2025** |  |  |  |
| Mean (SD) | 0.197 (0.595) | 0.853 (1.38) | 0.209 (0.626) |
| Median [Min, Max] | 0 [0, 15.0] | 0 [0, 14.0] | 0 [0, 15.0] |
| **Em Adm April 2025** |  |  |  |
| Mean (SD) | 0 (0) | 1.07 (0.299) | 0.0765 (0.288) |
| Median [Min, Max] | 0 [0, 0] | 1.00 [1.00, 4.00] | 0 [0, 4.00] |
| Missing | 84431 (75.2%) | 0 (0%) | 84431 (73.8%) |
| **Em Adm April 2025 To March 2026** |  |  |  |
| Mean (SD) | 0.227 (0.630) | 2.01 (1.58) | 0.260 (0.703) |
| Median [Min, Max] | 0 [0, 13.0] | 2.00 [1.00, 29.0] | 0 [0, 29.0] |

### Discrimination

Both models showed good discrimination for 1 month emergency admission (**Figure 2**). The Johns Hopkins ACG model achieved an AUC of 0.739 (95%CI 0.728 to 0.749), while ePRISM achieved 0.860 (95%CI: 0.852 to 0.867). The difference in AUC (0.121 [95%CI: 0.111 to 0.130]) was statistically significant by DeLong’s test (P< 0.0001), reflecting the large sample size and correlation between scores. In subgroup analyses for outcome at 1 month (**Supplemental Table 1**), the ePRISM model consistently outperformed Johns Hopkins ACG in both sexes and across all age bands (all p < 0.0001). AUCs were 0.857 (95%CI: 0.854 to 0.875) vs (95%CI:0.722 to 0.752) in females and 0.865 (95%CI:) vs 0.741 (95%CI: 0.724 to 0.755) in males; by age band, AUCs were 0.880 (95%CI: 0.868 to 0.891) vs 0.762 (95%CI: 0.743 to 0.780) (75–79 years), 0.857 (95%CI: 0.844 to 0.870) vs 0.726 (95%CI: 0.704 to 0.747) (80–84 years), and 0.816 (95%CI: 0.804 to 0.830) vs 0.676 (95%CI: 0.658 to 0.697) (≥85 years) for ePRISM versus Johns Hopkins ACG, respectively.

**Figure 2.**
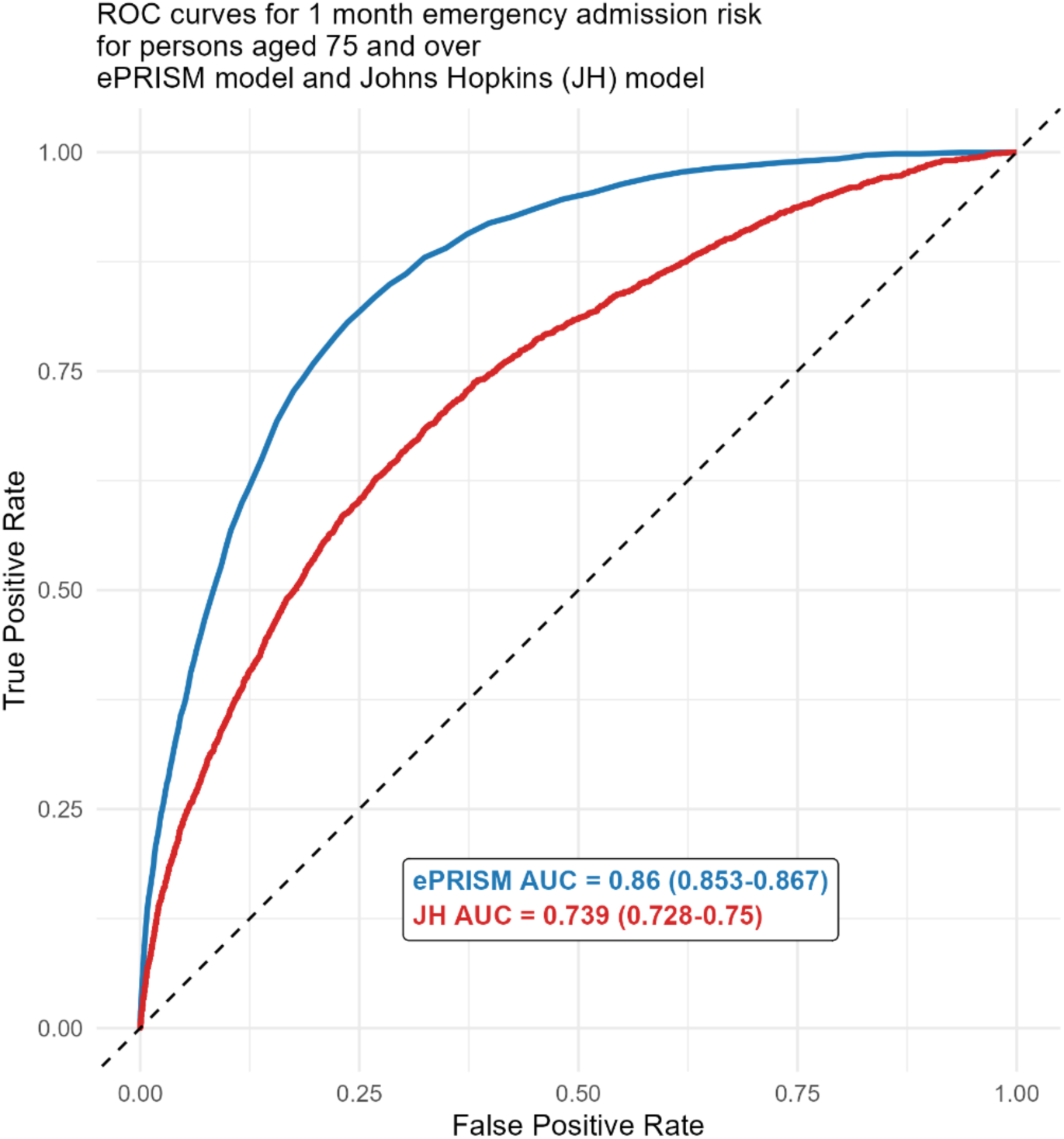
ROC curves for ePRISM and Johns Hopkins ACG

### Calibration and overall accuracy

Calibration assessment revealed more pronounced differences between the models. ePRISM exhibited relatively good calibration: observed event rates closely followed the identity line across most deciles of predicted risk, with only slight under-prediction at higher risk levels (**Figure 3; Supplemental Figures 1–2**). In keeping with this visual pattern, the calibration intercept and slope for ePRISM were close to their ideal values (intercept≈0; slope≈1), indicating that, on average, ePRISM risk estimates aligned well with observed event rates.

**Figure 3.**
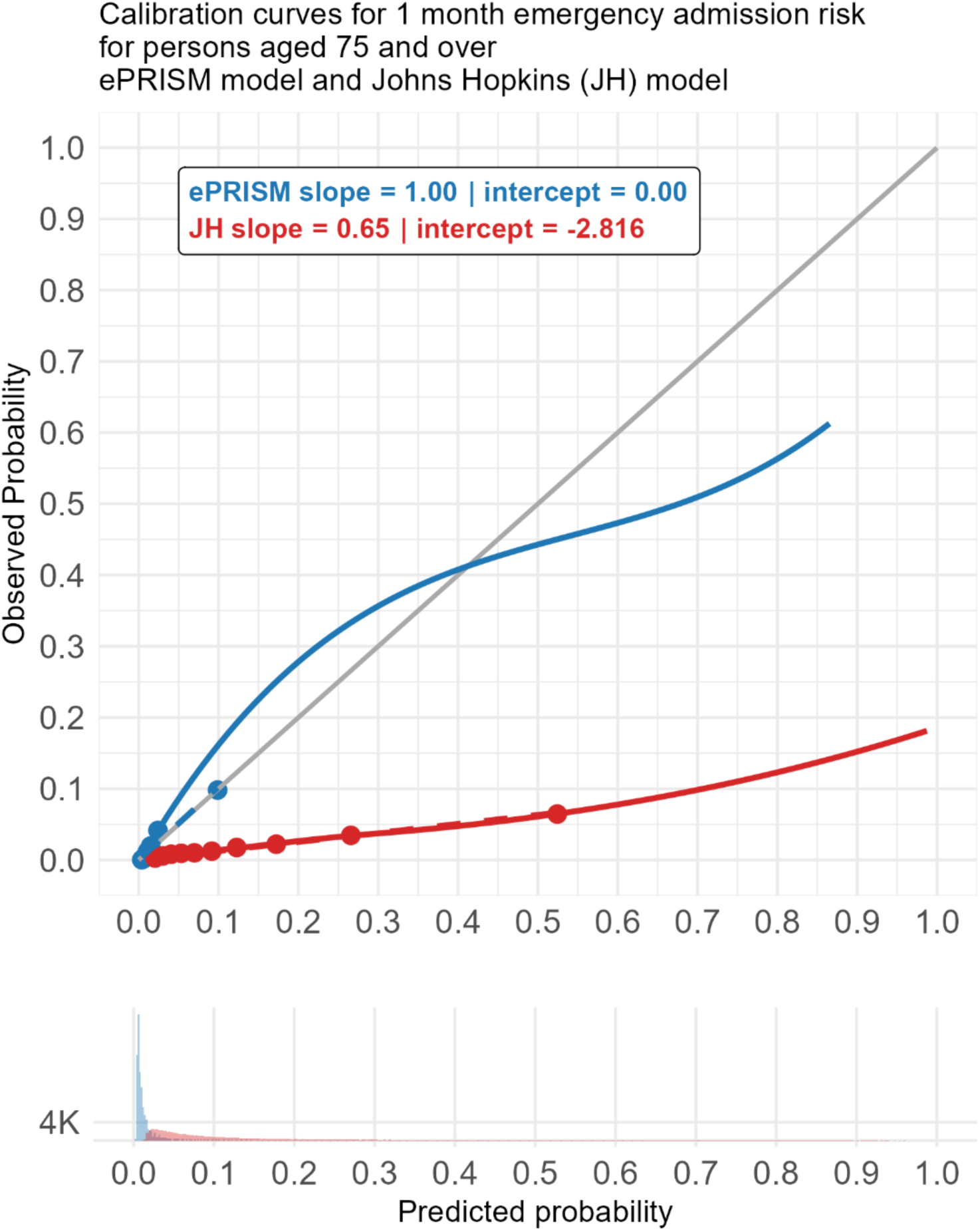
Calibration plots for ePRISM and Johns Hopkins ACG

In contrast, the Johns Hopkins ACG score showed systematic miscalibration, with predicted probabilities consistently higher than observed event rates across much of the spectrum. This pattern reflected systematic over-prediction of risk (**Figure 3; Supplemental Figures 1–2)**. Hosmer–Lemeshow statistics were statistically significant for both models; however, given the very large sample size, these results were interpreted descriptively and in conjunction with the graphical calibration patterns (**Figure 3**).

Brier scores highlighted the impact on overall accuracy: the Johns Hopkins model had a Brier score of 0.051 (95% CI 0.05 to 0.051), whereas ePRISM had a substantially lower Brier score of 0.017 (95% CI 0.017 to 0.018), indicating better overall predictive accuracy and reliability of probabilities for ePRISM **(Supplemental Table 2)**.

### Decision curve analysis

Decision curve analysis demonstrated that ePRISM delivered higher net benefit than both the “treat all” and “treat none” strategies across a range of plausible thresholds (**Supplemental Table 3**), and generally outperformed the Johns Hopkins model **(Figure 4**). In contrast, the Johns Hopkins ACG model yielded lower net benefit than ePRISM across most thresholds and fell below the “treat none” strategy across a wide portion of the threshold range, indicating potential harm (net benefit <0) if used to guide intervention at those thresholds (**Figure 3**). This pattern is consistent with the observed miscalibration of the Johns Hopkins score (**Figure 2**), which would increase false positives relative to true positives at commonly used decision thresholds.

**Figure 4.**
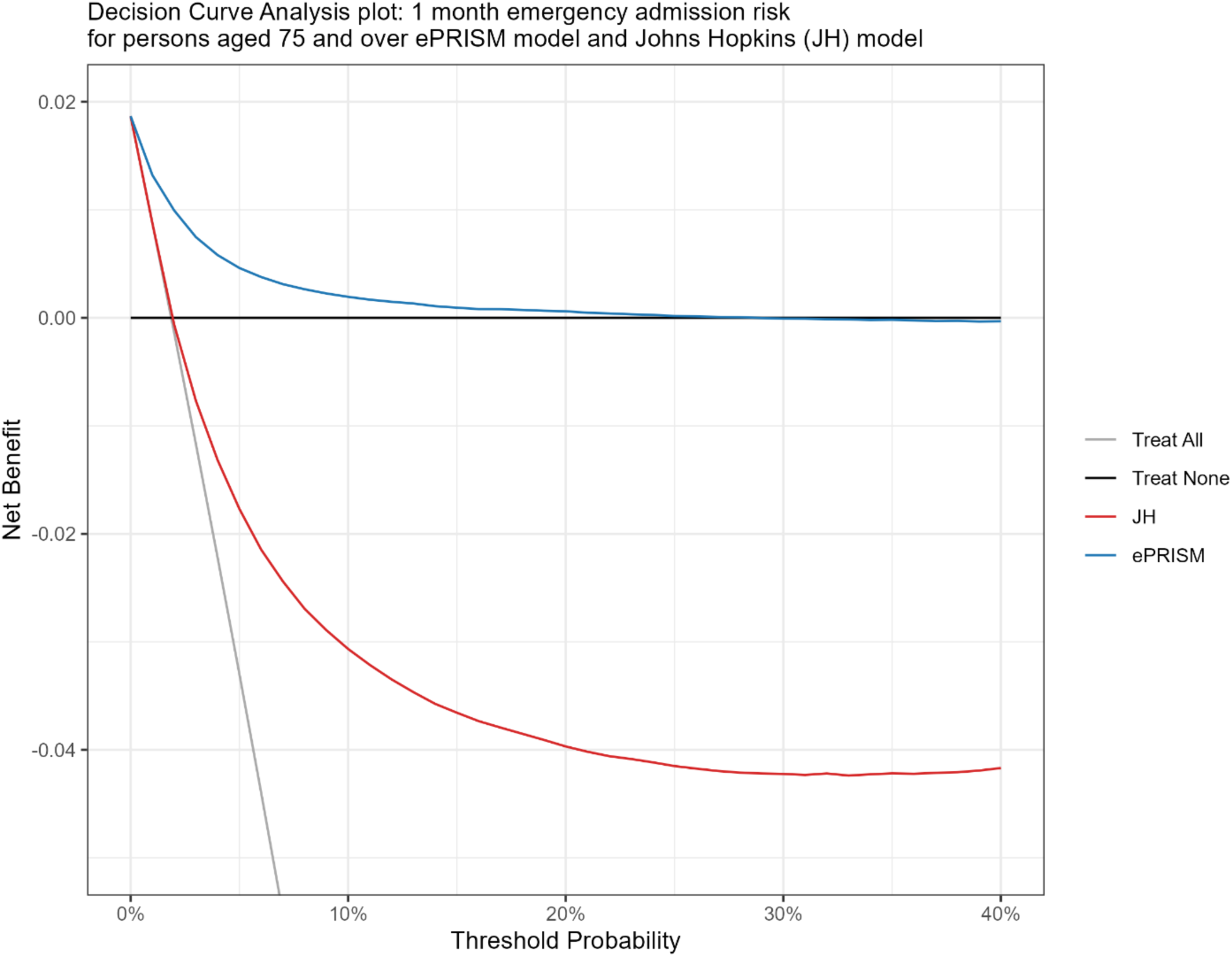
Decision curves for ePRISM and Johns Hopkins ACG

## Discussion

### Main findings

In this large cohort of 114,407 adults aged ≥75 years in an English ICS, ePRISM demonstrated higher discrimination than Johns Hopkins ACG for predicting 1-month emergency hospital admission. ePRISM also showed more favourable calibration, lower Brier score and higher decision-analytic net benefit across clinically plausible thresholds. Johns Hopkins ACG retained moderate discrimination, but its predicted probabilities were substantially higher than observed 1-month admission risks across much of the risk range, leading to poorer calibration and lower or negative net benefit at commonly considered decision thresholds. Taken together, these findings suggest that, in this local older population and for this short-term prediction horizon, ePRISM provided more reliable risk stratification for operational case-finding than Johns Hopkins ACG.

### Comparison with previous literature

Our results complement and extend the existing evidence base on emergency admission risk stratification tools. Kingston and colleagues reported that EARS tools are widely available in UK primary care, but that their implementation and use vary considerably and that evidence for improved patient outcomes remains limited [3].

Similarly, the PRISMATIC evaluation in Wales found that introducing PRISM for case-finding did not reduce emergency admissions and may increase contacts with health services when not embedded within clearly defined intervention pathways [5,21]. These studies highlight that predictive performance alone is not sufficient to improve outcomes, but robust local validation is still an essential first step before a score is used to target care.

Several studies have developed or validated prediction models for hospital admission in older or community-dwelling populations using routine primary care or administrative data, often reporting C-statistics around 0.7 or higher, although performance varies by population, data sources and outcome horizon [22,23]. The Johns Hopkins ACG result in our study is broadly consistent with this level of discrimination, whereas the ePRISM C-statistic for the 1-month outcome was higher. However, a key contribution of the present study is not only the comparison of AUCs but the simultaneous assessment of calibration, Brier score and decision-curve performance. Many earlier evaluations have emphasised rank-ordering ability, while probability calibration and clinical utility have been reported less consistently [8,18].

Guidance for commissioners has emphasised that no single predictive risk model is ideal and that performance may vary by local population, available data and intended use [24,7]. Our study contributes to this literature by providing a direct head-to-head comparison of two operational risk stratification tools applied to the same ICS population and evaluated against the same 1-month outcome. The finding that Johns Hopkins ACG, a well-established case-mix system, was less well calibrated than ePRISM in this older cohort reinforces the importance of local validation rather than assuming that a generic or externally supplied risk score will perform optimally in all settings [8,14].

Our emphasis on calibration and clinical utility aligns with TRIPOD and TRIPOD+AI recommendations and with broader methodological guidance for evaluating prediction models [8,17,18]. A model with good discrimination may still be unsuitable for threshold-based decision-making if its absolute probabilities are poorly calibrated [9]. This was evident in our decision curve analysis: Johns Hopkins ACG identified very large numbers of patients as high risk at common thresholds but generated a low positive predictive value and negative net benefit, whereas ePRISM identified a smaller and more enriched high-risk group. These findings demonstrate why calibration and decision-analytic assessment should be considered alongside AUC when selecting risk stratification tools for implementation [9,10].

### Clinical implications

For commissioners and clinicians in Norfolk and Waveney ICS, these findings have practical implications. Both ePRISM and Johns Hopkins ACG can rank older patients according to 1-month emergency admission risk, but the models differ materially in how many patients they would flag and how trustworthy their predicted probabilities appear to be. The overprediction observed for Johns Hopkins ACG could lead to excessive numbers of patients being classified as high risk, potentially diluting intervention resources, increasing workload and contributing to alert fatigue.

In contrast, ePRISM’s better calibration and positive net benefit suggest that its risk estimates may be more useful for short-term operational targeting. For example, at a 10% threshold, ePRISM flagged far fewer patients than Johns Hopkins ACG but achieved a higher positive predictive value and positive net benefit. This does not mean that ePRISM should be used mechanically at a single universal threshold. Rather, local teams should select thresholds in light of intervention capacity, the expected benefit of proactive care and the consequences of false positives and false negatives [9,10].

More broadly, our findings reinforce that discrimination alone is not sufficient for choosing between risk scores. ICSs and practices should request evidence on calibration plots, calibration intercept and slope, Brier scores and decision curves when evaluating commercial or locally implemented tools, particularly in older and multimorbid populations where baseline risk is high and miscalibration can have substantial consequences [8,9,10].

Finally, our results should be interpreted in the context of previous evidence showing that implementing risk stratification tools without clear clinical pathways and capacity for intervention is unlikely to reduce admissions and may even increase utilisation [5,6,21]. Choosing a better-calibrated model is therefore a necessary but not sufficient condition for impact. Risk stratification should be combined with well-designed intervention pathways, monitoring of downstream service use and prospective evaluation of patient and system outcomes [2,7].

## Strengths and limitations

A key strength of this study is the large, real-world cohort of 114,407 older adults within an ICS, providing precise estimates of performance for both models and allowing meaningful decision-analytic evaluation. The use of linked primary-care and hospital activity data reflects the information available to commissioners and mirrors how these tools are deployed in practice. The focus on adults aged ≥75 years addresses an important population for whom unplanned admissions are particularly consequential and for whom proactive care and case-finding are frequently advocated. Methodologically, we followed contemporary recommendations by examining discrimination, calibration, Brier scores and decision curves rather than relying on a single performance metric [8,17,18]. We also provided subgroup analyses by age and sex, which showed broadly consistent patterns.

Several limitations should be acknowledged. First, the analysis was conducted in a single ICS in England; performance and calibration may differ in other regions with different demographics, coding practices, service configurations or intervention pathways. Local validations elsewhere are therefore essential before generalising our findings. Second, both ePRISM and Johns Hopkins ACG are proprietary tools, and we did not have access to the full model specifications; this limited our ability to examine variable contributions, model updating or alternative recalibration approaches in depth. Third, the primary analysis focused on a 1-month prediction horizon, which aligns well with short-term operational risk stratification and the intended use of ePRISM, but performance may differ for longer horizons or when scores are updated repeatedly over time. Fourth, the prediction horizon and scaling of the Johns Hopkins ACG probability should be considered when interpreting calibration, Brier score and decision-curve results for a 1-month outcome; locally recalibrated probabilities may be required before using either score for threshold-based decisions. Fifth, our decision curve analysis summarises clinical utility using net benefit, but the consequences of false positives and false negatives will vary by local context and intervention type. Finally, we evaluated predictive performance only; we did not assess whether using ePRISM or Johns Hopkins ACG to trigger interventions improves patient outcomes or reduces resource use. Implementation studies will be needed to determine how best to integrate risk tools into care pathways.

## Conclusions

In this external validation and comparative study of two emergency admission risk scores in older adults, ePRISM demonstrated higher discrimination than Johns Hopkins ACG for predicting 1-month emergency admission. ePRISM also showed more favourable calibration, lower Brier score and greater decision-analytic net benefit, indicating better apparent suitability for short-term risk-based targeting in the Norfolk and Waveney ICS population.

Our findings underscore that model selection should not be based on AUC alone. Calibration and decision-analytic measures are crucial when choosing between competing risk scores, particularly for high-risk older populations where miscalibration can lead to misallocation of scarce preventive resources. In this ICS and for this 1-month prediction horizon, ePRISM appears to be the more appropriate tool for targeting proactive care in older adults, while ongoing monitoring, recalibration where necessary and prospective evaluation of implementation impact remain important.

## List of abbreviations

ACG: Adjusted Clinical Groups
AUC: Area under the receiver operating characteristic curve
DCA: Decision curve analysis
EARS: Emergency Admission Risk Stratification
HES: Hospital Episode Statistics
ICS: Integrated Care System
NHS: National Health Service
PRISM: Predictive Risk Stratification Model
ROC: Receiver operating characteristic
TRIPOD: Transparent Reporting of a multivariable prediction model for Individual Prognosis Or Diagnosis

## Declarations

### Ethics approval and consent to participate

This study involved secondary analysis of pseudonymised routinely collected health-care data. Ethical approval was obtained from the South West - Exeter Research Ethics Committee (REC reference: 17/SW/0001). Individual patient consent was not required because data were anonymised prior to analysis in line with national governance arrangements.

### Consent for publication

Not applicable.

### Availability of data and materials

The datasets analysed are not publicly available as they contain individual-level health-care data subject to NHS data-sharing agreements.

## Competing interests

The authors declare that they have no competing interests.

## Funding

None.

## Authors’ contributions

*DY* conceived the study and led the analysis. TW and AP extracted and curated the data. TW and AP conducted the statistical analyses with input from *DY*. All authors contributed to interpretation of findings. *DY*drafted the manuscript, and all authors critically reviewed and approved the final version.

## Acknowledgements

We thank colleagues in Norfolk and Waveney ICS, the Eclipse and commissioning support teams for providing data access and technical support.

---

**Supplemental Figure 1.**
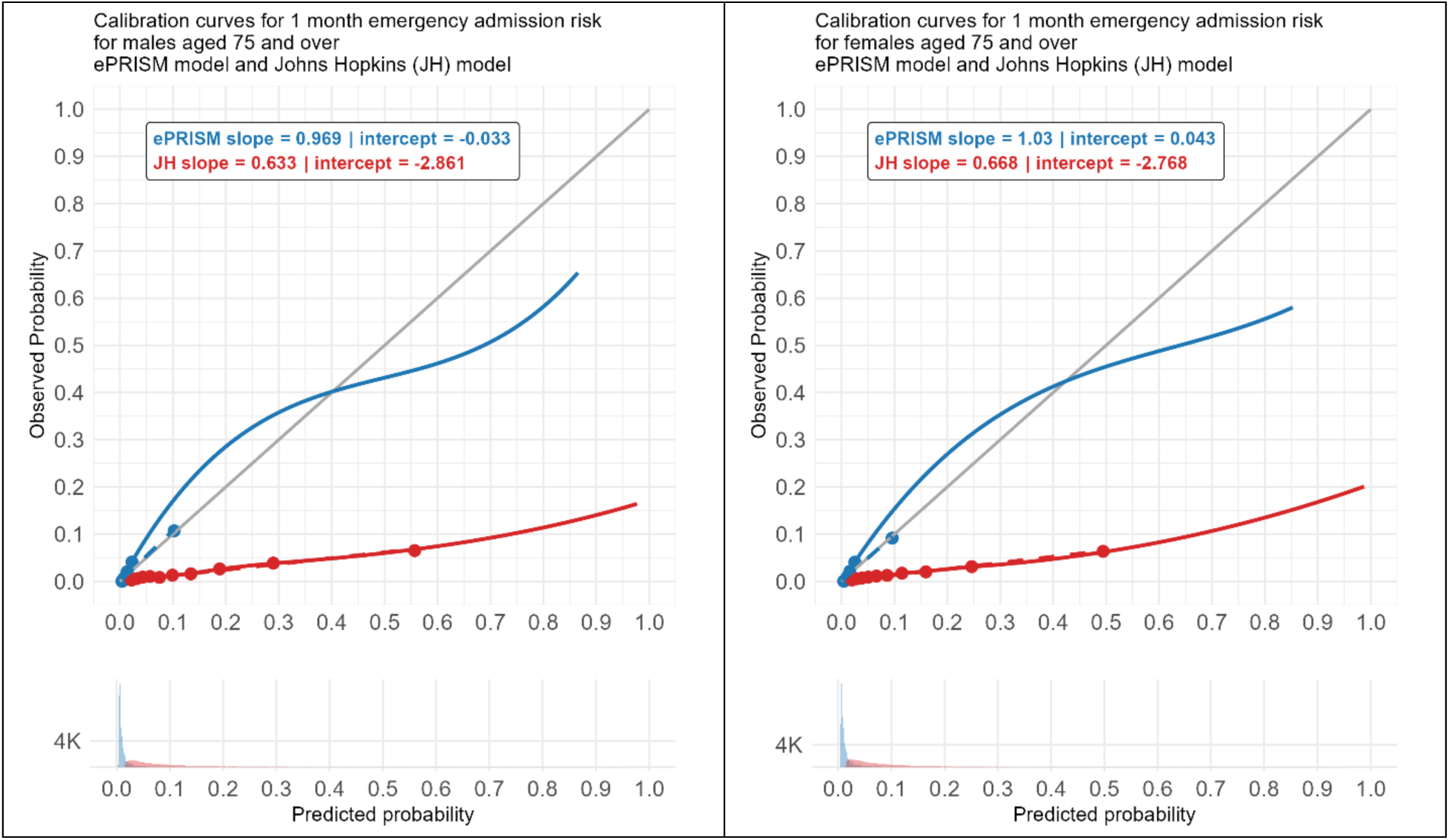
Calibration plots for ePRISM and Johns Hopkins ACG by gender

**Supplemental Figure 2.**
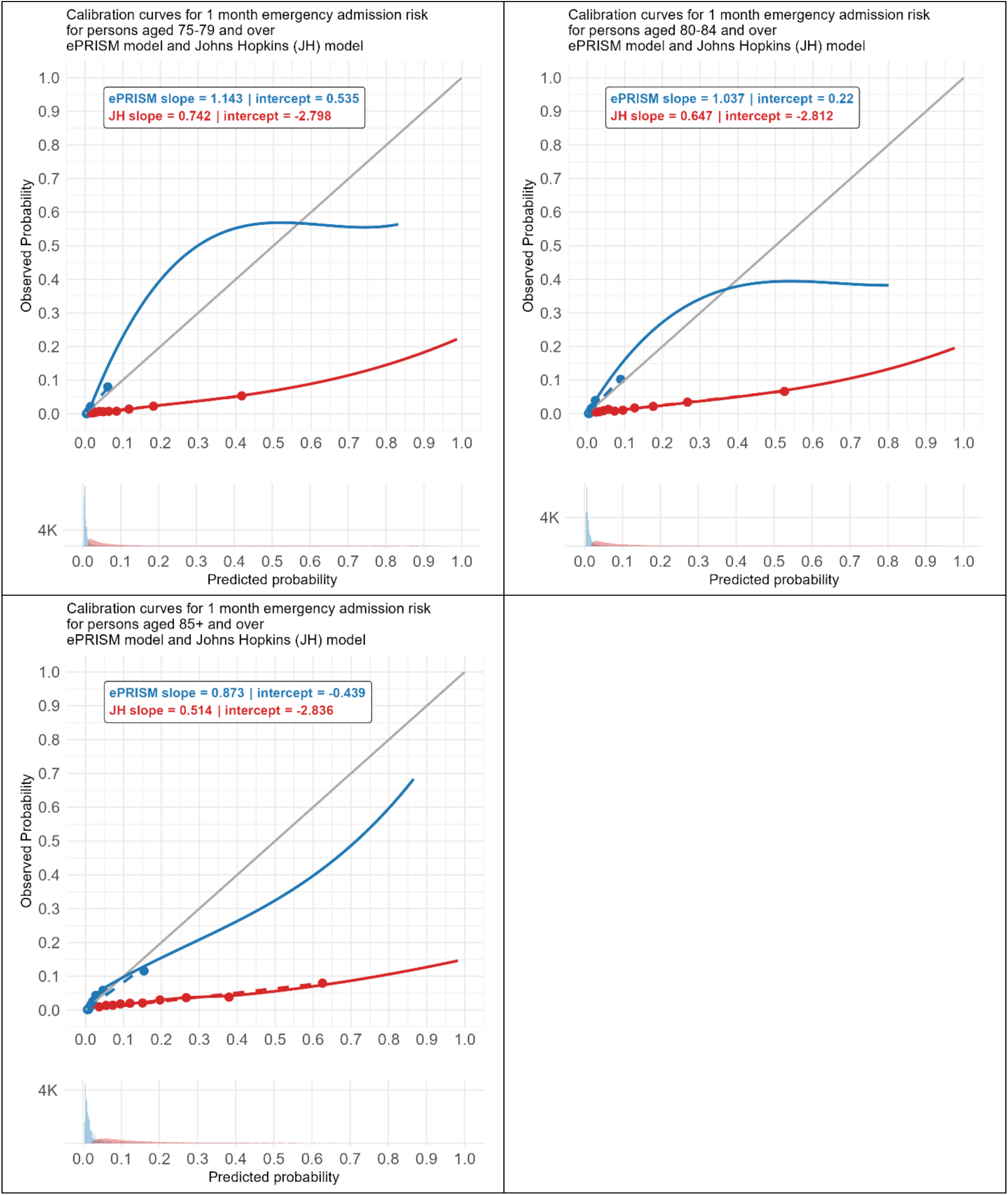
Calibration plots for ePRISM and Johns Hopkins ACG by age group

**Supplemental Table 1.** C-statistics for ePRISM and Johns Hopkins ACG in predicting 1-month emergency admission risk by subgroup. DeLong test was used to compare the C-statistics between two risk scores.

| Age Band | Sex | C-statistic (95% CI)<br>for Johns Hopkins<br>ACG score | C-statistic (95% CI)<br>for ePRISM | C-statistics difference<br>(95%CI) | *P*-values |
| --- | --- | --- | --- | --- | --- |
| 75+ | All | 0.739 (0.728 to 0.749) | 0.860 (0.852 to 0.867) | 0.121(0.111 to 0.13) | *P* < 0.00001 |
| 75+ | Male | 0.741 (0.724 to 0.755) | 0.865 (0.854 to 0.875) | 0.124 (0.111 to 0.137) | *P* < 0.00001 |
| 75+ | Female | 0.737 (0.722 to 0.752) | 0.857 (0.847 to 0.866) | 0.120 (0.107 to 0.132) | *P* < 0.00001 |
| 75-79 | All | 0.762 (0.743 to 0.78) | 0.880 (0.868 to 0.891) | 0.119 (0.113 to 0.15) | *P* < 0.00001 |
| 80-84 | All | 0.726 (0.704 to 0.747) | 0.857 (0.844 to 0.87) | 0.132 (0.113 to 0.15) | *P* < 0.00001 |
| 85+ | All | 0.676 (0.658 to 0.697) | 0.816 (0.804 to 0.83) | 0.140 (0.123 to 0.157) | *P* < 0.00001 |

**Supplemental Table 2.**
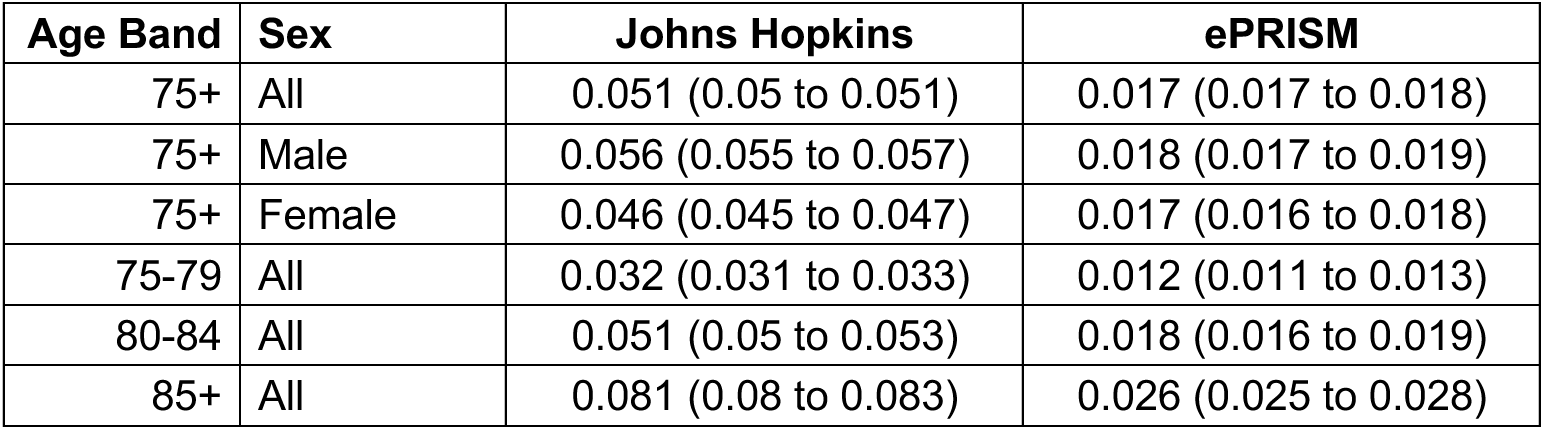
Brier Score (95% CI) for ePRISM and Johns Hopkins ACG in predicting 1-month emergency admission risk by subgroup.

**Supplemental Table 3.** Net Benefit at selected thresholds for decision curve analysis.

| Model | Threshold | Number at high risk | Sensitivity | Specificity | PPV | True +ve | False +ve | True -ve | False -ve | Net Benefit |
| --- | --- | --- | --- | --- | --- | --- | --- | --- | --- | --- |
| JH | 0.05 | 76917 | 0.900 | 0.332 | 0.025 | 1923 | 74994 | 37277 | 213 | -0.018 |
| JH | 0.10 | 47732 | 0.757 | 0.589 | 0.034 | 1616 | 46116 | 66155 | 520 | -0.031 |
| JH | 0.15 | 32764 | 0.636 | 0.720 | 0.041 | 1359 | 31405 | 80866 | 777 | -0.037 |
| JH | 0.20 | 23971 | 0.544 | 0.797 | 0.048 | 1162 | 22809 | 89462 | 974 | -0.040 |
| JH | 0.25 | 18188 | 0.462 | 0.847 | 0.054 | 986 | 17202 | 95069 | 1150 | -0.042 |
| JH | 0.30 | 14089 | 0.396 | 0.882 | 0.060 | 845 | 13244 | 99027 | 1291 | -0.042 |
| ePRISM | 0.05 | 7351 | 0.407 | 0.942 | 0.118 | 869 | 6482 | 105789 | 1267 | 0.005 |
| ePRISM | 0.10 | 3319 | 0.249 | 0.975 | 0.160 | 532 | 2787 | 109484 | 1604 | 0.002 |
| ePRISM | 0.15 | 1857 | 0.173 | 0.987 | 0.199 | 370 | 1487 | 110784 | 1766 | 0.001 |
| ePRISM | 0.20 | 1097 | 0.129 | 0.993 | 0.251 | 275 | 822 | 111449 | 1861 | 0.001 |
| ePRISM | 0.25 | 767 | 0.096 | 0.995 | 0.269 | 206 | 561 | 111710 | 1930 | 0.000 |
| ePRISM | 0.30 | 524 | 0.071 | 0.997 | 0.288 | 151 | 373 | 111898 | 1985 | 0.000 |

**Supplemental Table 4.**
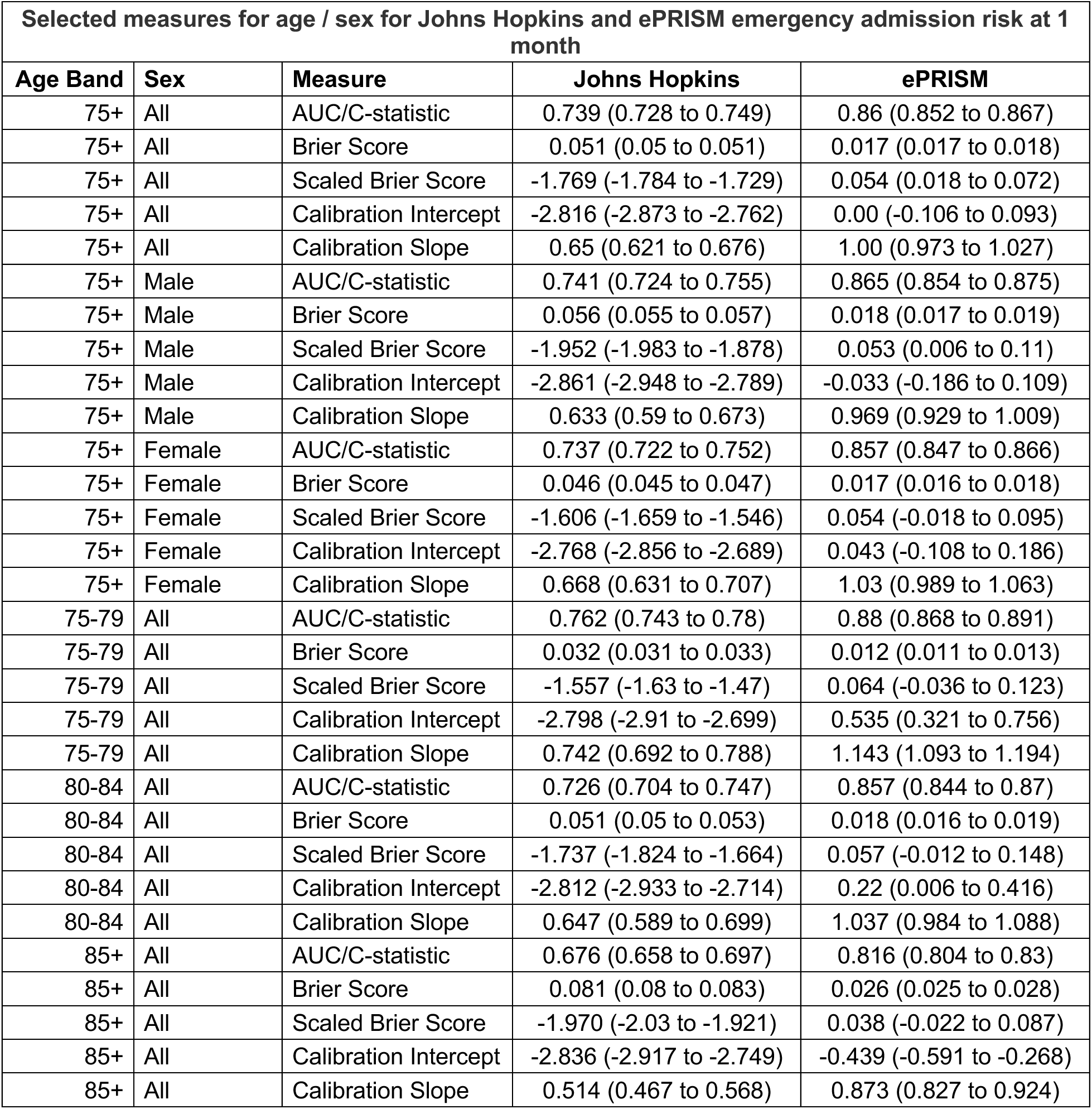
Selected measures by age and sex for comparing Johns Hopkins and ePRISM models for emergency admission at 1 month.

## Notes

### Competing Interest Statement

The authors have declared no competing interest.

### Author Declarations

This study involved secondary analysis of pseudonymised routinely collected health-care data. Ethical approval was obtained from the South West ‐ Exeter Research Ethics Committee (REC reference: 17/SW/0001). Individual patient consent was not required because data were anonymised prior to analysis in line with national governance arrangements.

